# Coupling freedom from disease principles and early warning from wastewater surveillance to improve health security

**DOI:** 10.1101/2021.06.11.21258797

**Authors:** D. A. Larsen, M. B. Collins, Q. Du, D. Hill, T. Z. Insaf, P. Kilaru, B. L. Kmush, F. Middleton, A. Stamm, M. L. Wilder, T. Zeng, H. Green

## Abstract

Infectious disease surveillance is vitally important to maintaining health security, but these efforts are challenged by the pace at which new pathogens emerge. Wastewater surveillance can rapidly obtain population-level estimates of disease transmission, and we leverage freedom from disease principles to make use of non-detection of SARS-CoV-2 in wastewater to estimate the probability that a community is free from SARS-CoV-2 transmission. From wastewater surveillance of 24 treatment plants across upstate New York beginning in May 2020, we observed a reliable limit of detection of 0.3--0.5 cases per 10,000 population. No COVID-19 cases were reported 40% of the time following a non-detection of SARS-CoV-2 in wastewater, and cases were less than 1 daily case per 10,000 population 97% of the time following non-detection. Trends in the intensity of SARS-CoV-2 in wastewater correlate with trends in COVID-19 incidence and test positivity (ρ>0.5), with the greatest correlation observed for active cases and a three-day lead time between wastewater sample date and clinical test date. Wastewater surveillance can cost-effectively demonstrate the geographic extent of the transmission of emerging pathogens, confirming that transmission is absent or under control and alerting of an increase in transmission. If a statewide wastewater surveillance platform had been in place prior to the onset of the COVID-19 pandemic, policymakers would have been able to complement the representative nature of wastewater samples to individual testing, likely resulting in more precise public health interventions and policies.

---

Infectious disease surveillance, or monitoring trends in the transmission of communicable diseases, is an important pillar of public health and is fundamental to the health security of a population.^1–3^ Infectious disease surveillance systems are typically event-based, designed to measure abnormal increases in either identified cases of a laboratory-diagnosed pathogen or identified cases where individuals display a set of signs and symptoms (syndromic surveillance).^4^ Although infectious disease surveillance is necessary to identify and control disease outbreaks,^5^ novel pathogens are incredibly difficult to incorporate into existing event-based surveillance networks. For example, by the time that sudden acute respiratory coronavirus 2 (SARS-CoV-2) was first noticed via an increase of unexplainable cases of pneumonia,^6^ the pathogen had spread widely through the community in Wuhan, China. Subsequently an event-based surveillance approach was not a readily feasible strategy to confirm that connected communities were free from SARS-CoV-2 transmission as the pathogen spread across the globe. Despite heroic efforts to scale COVID-19 diagnostics as quickly as possible, the virus spread more rapidly, exceeding testing capacity early in the pandemic. Without an operational event-based surveillance strategy based upon virus diagnostics, officials in the US and elsewhere had to rely on syndromic surveillance as SARS-CoV-2 spread--basing public health interventions on increases in hospitalizations and deaths. Since timely, geographically granular information regarding transmission was not available, public health officials were left with few management options, other than to call for a halt to non-essential businesses and gatherings. This type of broad, country-wide reaction highlights an inadequate infectious disease surveillance system that could not provide reliable information on where SARS-CoV-2 transmission was, and more importantly, where transmission was not occurring.^7^ Such information, especially regarding communities that were actually free from transmission at the time, would have allowed for local decision making and precise interventions in response to the arrival of SARS-CoV-2 into the community.^8,9^ Instead, the unknown spread of the virus forced statewide decisions, similar to many states across the country, ending in-person schooling and shuttering non-essential businesses statewide.^10^

Wastewater surveillance addresses challenges and limitations of event-based surveillance systems by assessing infectious disease transmission potential within the entire population served by a sewage system. A single sample conveys information independent of individuals’ symptoms, health-seeking behavior, or the health system’s availability of diagnostic testing.^11^ The genetic material from SARS-CoV-2 (as with many infectious pathogens) is shed in human feces and urine.^12–14^ Finding that genetic material in wastewater can serve as an early indication of a pathogen in a community, as demonstrated in the Netherlands, Paris, Connecticut, and elsewhere.^15–18^ Perhaps more importantly, absence of genetic material in the wastewater can give confidence that a community is free from transmission of that pathogen. Tracking infectious disease transmission through wastewater is not a novel approach - it goes back more than 100 years^19–21^ and saw broadscale application in the later 1900’s to identify outbreaks of polio.^22^ Wastewater surveillance in some cases even allowed for vaccine campaigns to control community polio outbreaks before any polio-caused paralysis occurred.^23,24^ Beginning in May of 2020, we analyzed wastewater from 76 sewer sheds and 24 wastewater treatment plants in 14 counties throughout upstate New York (Supplemental Fig. 1). Here we compare levels of SARS-CoV-2 RNA found in the wastewater to measures of SARS-CoV-2 transmission as reported by the New York State Department of Health. Using freedom from disease principles,^25^ we then assessed the ability of wastewater surveillance to confirm that SARS-CoV-2 transmission was either under control or absent. By comparing spikes in SARS-CoV-2 as measured in wastewater to spikes in event-based surveillance measures of SARS-CoV-2 transmission we determined whether wastewater surveillance provided early warning of localized outbreaks in three communities. Finally, we estimated the cost of a statewide wastewater surveillance platform for NY State. Together, these analyses provide compelling evidence that a wastewater surveillance network would be a cost-effective system to increase health security.

## Leveraging wastewater surveillance data to confirm freedom from disease

We can leverage negative results from wastewater surveillance to establish the probability that SARS-CoV-2 transmission is below a measurable and manageable threshold using principles of freedom from disease surveillance developed by veterinary scientists.^25^ Each non-detection of SARS-CoV-2 in wastewater carries one of two possibilities (Fig. 1). Either SARS-CoV-2 is absent from the population or the wastewater test was unable to detect SARS-CoV-2. The ability of wastewater to detect SARS-CoV-2, or the sensitivity of the wastewater surveillance approach in this context, is a function of the method’s limits of detection, the population size of the sewershed, dilution of human waste with precipitation and graywater, and the proportion of the population in the sewershed with sewer connections. Using our novel ultracentrifugation method,^26^ we observed detection of SARS-CoV-2 RNA from a 24-hour composite sample of wastewater to be between 0.3--0.5 daily cases per 10,000 people or 0.5--1% test positivity (all connected to sewers) one day following the wastewater sample (Fig. 2). At that time, 40% of the 472 sewershed time points with non-detection of SARS-CoV-2 RNA had zero incident cases, and 96% had fewer than one daily case per 10,000 population (Fig. 2). Sewersheds where there were COVID-19 cases reported but no detectable SARS-CoV-2 RNA in wastewater were, on average, five times more populous than sewersheds where SARS-CoV-2 RNA went undetected and no COVID-19 case was reported. The number of diagnostic tests conducted in these more populous sewersheds was slightly higher (a median of 12 compared to 9 per 1,000 population).

**Figure 1:**
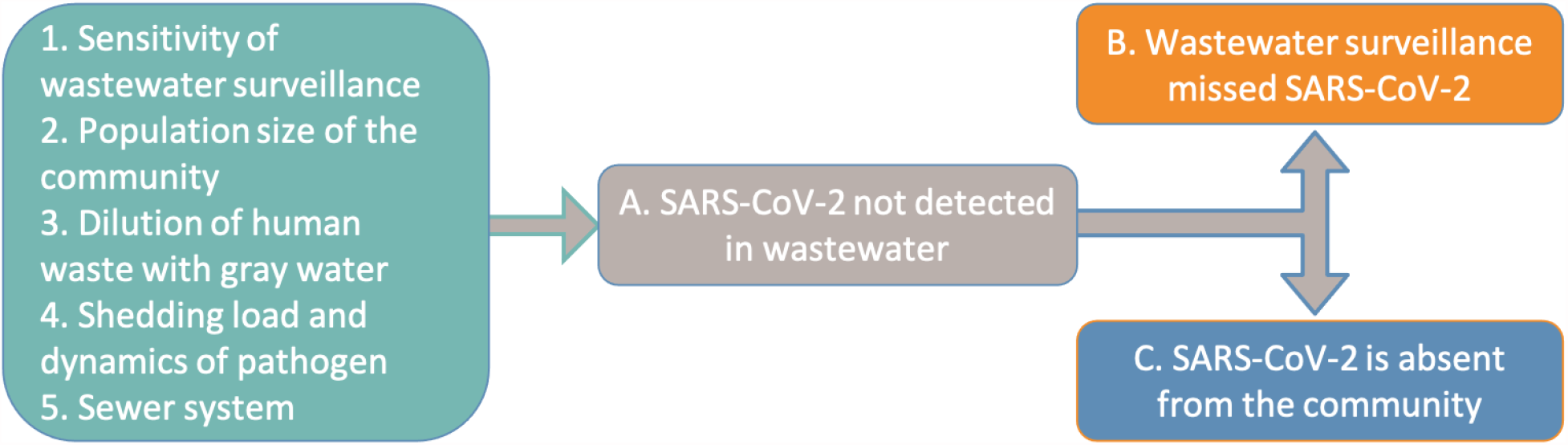
Each time wastewater is tested and SARS-CoV-2 is not detected (A), either the system did not detect the SARS-CoV-2 that was in the community (B) or there was no SARS-CoV-2 in the community (C). The probability of each non-detection representing no SARS-CoV-2 in the community is dependent upon the sensitivity of wastewater surveillance (1), the population size of the community (2), the dilution of human waste with gray water (3), the load shedding dynamics of the pathogen (4), and the sewer system (5). Whereas each non-detection may only give a low probability that SARS-CoV-2 is absent, repeatedly not detecting SARS-CoV-2 increases the confidence that SARS-CoV-2 is absent using the simple equation of 1-(1-sensitivity)^n^ where n represents the number of consecutive non-detections. This process is similar to repeatedly tossing a coin and repeatedly getting heads - each individual coin toss maintains a specific probability but the probability of obtaining a string of repeated consecutive results is increasingly lower with each toss.

**Figure 2:**
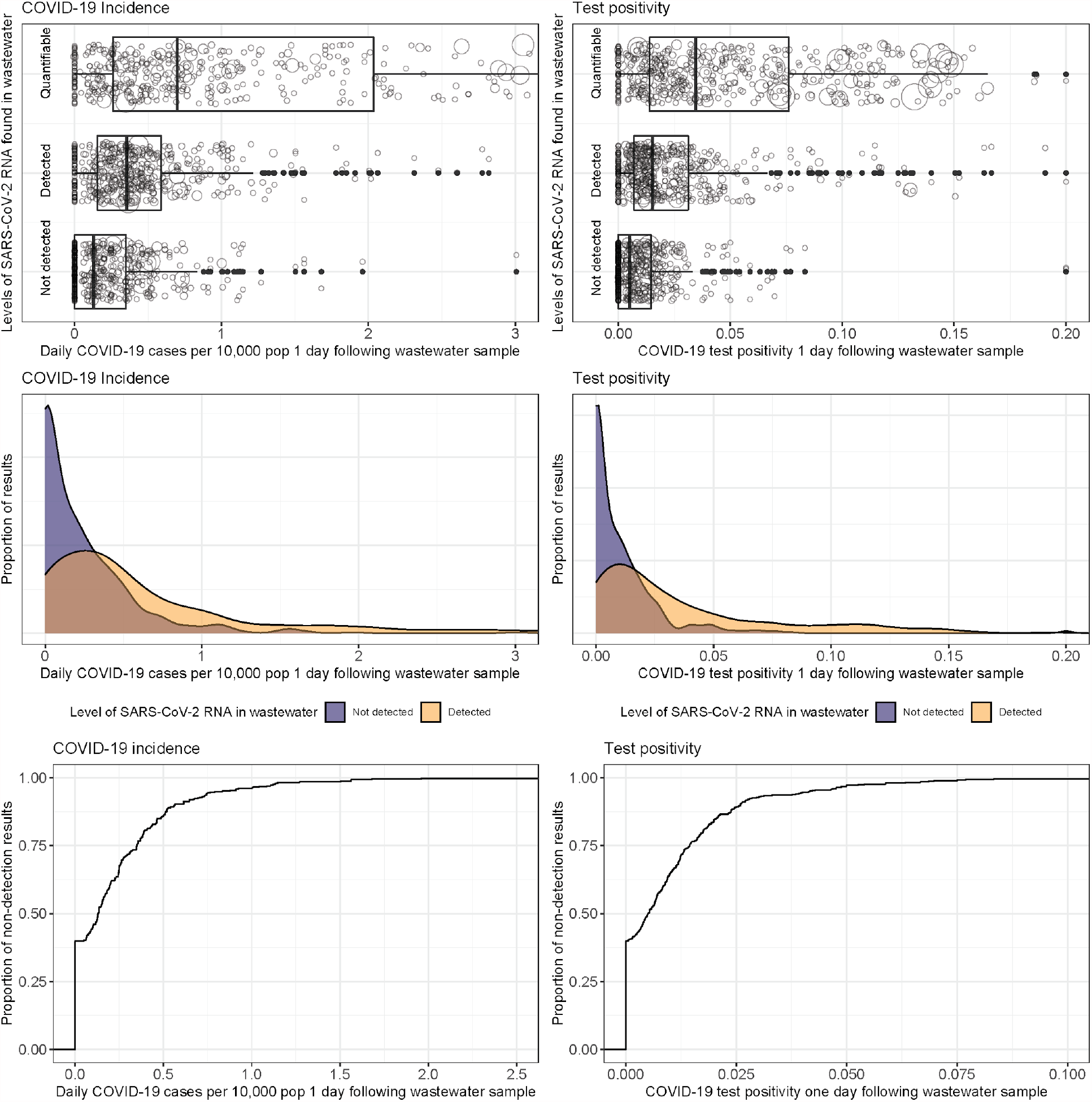
Estimates of the limits of detection (in terms of cases reported in the health system) of SARS-CoV-2 testing in wastewater. Clear differentiation in the level of measured community-level COVID-19 incidence (A) and test positivity (B) when categorizing wastewater results as quantifiable, detected but below the level of quantification, and not detected. Size of the circles in A, B represent the number of individuals tested. Large overlap between detection and nondetection exists (C, D), with non-detection of SARS-CoV-2 RNA clustering < 0.5 cases per 10,000 and < 2% test positivity (E, F)

By combining negative results (SARS-CoV-2 RNA undetected) over time from wastewater surveillance, we can estimate the probability that a community is free from coronavirus transmission, or at least that transmission is extremely low (less than 1 incident case per 10,000 population). Importantly, non-detections in larger sewersheds result in less confidence than smaller sewersheds. Among the sewersheds in our study, the potential sensitivity of a single non-detected sample ranged from 0.008 in the largest sewershed to 0.40 in smaller sewersheds for absence of infection and 0.02 to 0.97 for extremely low transmission (< 1 daily case per 10,000 population). Repeated wastewater samples where SARS-CoV-2 RNA is repeatedly undetected indicate an increased probability that the upstream community is either free from SARS-CoV-2 transmission or has it under control. This occurred in Cayuga and Cortland counties during the summer of 2020 (Supplemental Figure 2). While individuals in these counties were still at risk of contracting COVID-19 at this time, risk was more dependent on interaction with people outside the community (including recently returned travelers) than interaction with people inside the community. These findings would thus have allowed for social distancing interventions to be more precisely applied.

## Correlation between wastewater results and pandemic trajectory

We observed broad correlation (ρ∼0.5) between SARS-CoV-2 intensity in wastewater and COVID-19 case and testing data (Figure 3, Supplemental Figure 3), including a ρ=0.56 for active cases and ρ=0.55 for seven-day average test positivity with a three-day lead between wastewater sample and COVID-19 case data. For incident COVID-19 cases we observed a correlation of ρ = 0.55 with a six-day lag between wastewater sample and COVID-19 case data.

**Figure 3:**
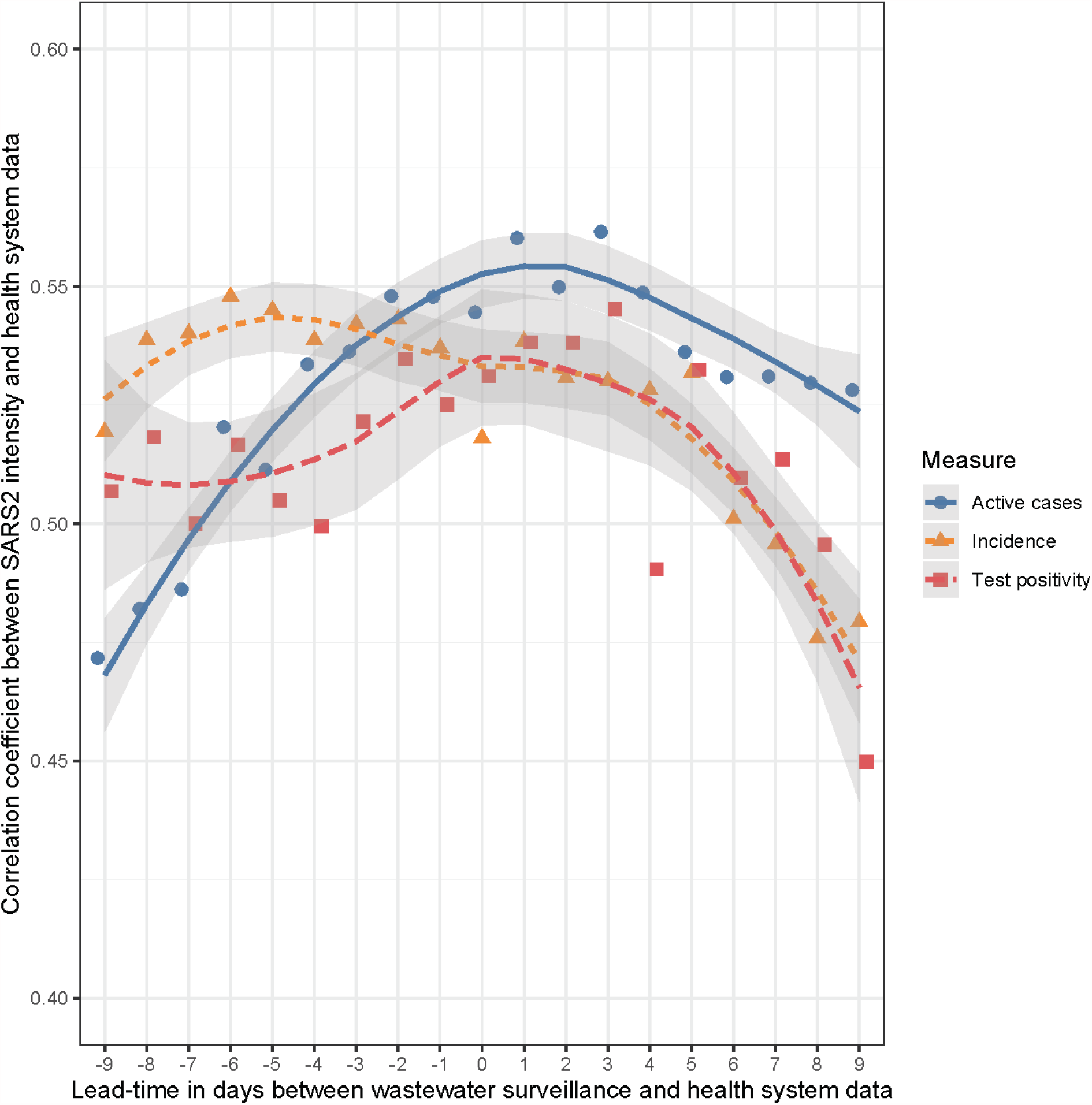
Correlation observed between intensity of SARS-CoV-2 RNA in wastewater and various indicators of SARS-CoV-2 transmission at different lag and lead times, with a locally weighted smoothing (lines) and uncertainty (gray shading).

## Wastewater surveillance provides early indication of increasing transmission

In the fall of 2020, three upstate New York communities monitoring coronavirus in their wastewater reported COVID-19 outbreaks following the return of college students to campus: Cortland, Oneonta, and Oswego. Prior to the beginning of the semester, in late August 2020, SARS-CoV-2 transmission was stable and low with an average daily incidence between July 1, 2020 and August 15, 2020 of less than 0.2 reported COVID-19 cases per 10,000 population. Reported COVID-19 cases increased sharply, beginning August 31 in Oneonta and September 10 in both Cortland and Oswego (Supplemental Fig 4). Following a long period of non-detection, SARS-CoV-2 RNA in Cortland wastewater samples was first detected on September 1 and increased to quantifiable levels on September 8 with a continued increase in quantifiable levels thereafter. Following a period of variable detection, but below the level of quantification, SARS-CoV-2 RNA in Oswego wastewater samples was first quantifiable on September 2, with increasing levels thereafter. In Oneonta, quantifiable levels were first observed September 1 and increased thereafter. Together these signals provided about a week’s warning of the three outbreaks observed (Supplemental Fig 4).

## Discussion

The primary public health benefit of wastewater surveillance resides in confirming that a community is safe from a pathogen, both through increased confidence in the absence of transmission and the early warning of a pathogen’s presence in a community. As a pathogen threatens a community, and particularly when human diagnostics are limited, this benefit is invaluable. These primary benefits associated with wastewater surveillance are dependent upon methodological limits of detection. In line with reports from Australia,^27^ we observed inaccuracy 60% of the time, that being when there is a mismatch between non-detection of SARS-CoV-2 RNA and reported COVID-19 cases in a sewershed. Despite what may appear to be a high rate of inaccuracy, 40% sensitivity in these types of surveillance systems is quite good, as is 96% sensitivity in a surveillance system to confirm that transmission is below 1 daily case per 10,000 population. To put these sensitivity estimates in context, in March of 2020 the health system was only about 10% sensitive to COVID-19 cases since as many as 90% of cases of COVID-19 in New York went undiagnosed.^28^ Additionally, wastewater surveillance is unaffected by health access disparities that lead to underreporting in vulnerable communities.^29^

We observed the highest correlation between levels of SARS-CoV-2 RNA in wastewater and health system measures of transmission with a 3-day lead time, similar to Peccia et al.^16^ Further, we saw greater correlation between active cases and wastewater than we observed with incident cases, suggesting that modelers should take post-infection shedding into account.

Wastewater surveillance will not capture all infections in a community, particularly in very large communities (> 50,000 population) and communities with a high proportion of houses that are not connected to the sewer. Fortunately for public health, single infections are less concerning than population-level trends (i.e. spread). With COVID-19 and many other infectious diseases, a majority of infections are dead-end transmission.^30,31^ Communities where houses are not connected to sewers would be inherently excluded from a wastewater surveillance program, but we would expect a pathogen to spread between both non-sewered and sewered households. Furthermore, when caseloads are low (daily cases < 1 per 10,000 population) the test, trace, isolate paradigm should be sufficient for population control.^32,33^ Rather than needing to find every instance of a single COVID-19 case in a community, the public and policy makers need to know when a pathogen first arrives in a community and when transmission is increasing beyond chains of known transmission to a state of community spread.

New York State’s health security was breached in 2020 with the undetected arrival and subsequent spread of SARS-CoV-2. The pandemic required broadscale closure of schools, businesses, and general social movement to “flatten the curve” in order to mitigate the surge in hospitalizations and death.^34^ The only reliable understanding of SARS-CoV-2 transmission at the time of intervention came via hospitalizations that were threatening to overwhelm and in some cases did overwhelm the health system. Interventions were implemented due to the potential of what could happen statewide rather than what was actually happening locally. Having a wastewater surveillance system in the face of such a public health emergency would have provided a more complete understanding of the geographic extent of SARS-CoV-2 transmission and resulting public health interventions could have been more precise and perhaps shorter. From our findings, wastewater surveillance as applied in upstate New York exceeds the need in this regard and should be considered an important aspect of a community’s health security.

## Data Availability

Wastewater data is available from the authors upon approval from the municipal wastewater treatment plants. COVID-19 case data is available from the New York State Department of Health following formal request and approval. All R scripts used in the analyses are available from the authors upon reasonable request.

## Acknowledgements

This work was made possible by participating wastewater treatment plants and their operators, especially those from Onondaga County who guided sampling and other key aspects of this work. The NY state pilot study was conducted in collaboration with Arcadis and Quadrant Biosciences.

## Author contributions

conceptualization - DAL, BLK, TZ, HG; data curation - DAL, MBC, DH, TI, AS; formal analysis - DAL, MBC, DH; funding acquisition - DAL, PK, HG; investigation - QD, FM, MW, HG; methodology - FM, MW, HG; project administration - DAL; resources - FM, HG; software - DAL, MBC; supervision - DAL, MBC, HG; validation - FM, MW, HG; visualization - DAL; writing - original draft - DAL; writing - review and editing - DAL, MBC, QD, DH, TI, PK, BLK, FM, AS, MW, TZ, HG.

## Supplementary Materials

### Materials and Methods

#### Wastewater testing

Participating wastewater treatment plants, or Arcadis during the New York State pilot, sent a 24-hour composite of 250mL untreated wastewater to Quadrant Biosciences either weekly, twice weekly, or three times weekly depending upon services contracted. Quadrant Biosciences purified and quantified SARS-CoV-2 viral nucleic acid levels contained in wastewater samples by using the ultracentrifugation through sucrose cushion (UltraSucrose) technique followed by qRT-PCR as detailed elsewhere.^26^ Briefly, wastewater was added to a centrifuge tube before adding sucrose solution under the wastewater creating two distinct layers. After ultracentrifugation, the supernatant was removed and the pellet containing nucleic acids was resuspended. Total nucleic acids were extracted from eluted pellets and used immediately as the template for quantification of SARS-CoV-2 and crAssphage DNA and crAssphage RNA.The following equation was used to normalize SARS-CoV-2 quantities to the level of fecal material in each sample as indicated by crAssphage DNA concentrations: log_10_(SARS-CoV-2):log10(crAssphage DNA).

#### COVID-19 case data

COVID-19 case data was pulled from the Electronic clinical Laboratory Reporting System (ECLRS). Every licensed professional authorized by the Department of Health Physician Office Laboratory Evaluation Program to administer a test for COVID-19 or influenza is required to report such results immediately (not more than 3 hours) to the Department of Health through ECLRS when a result is received.^10,35^ COVID-19 cases and tests were retrieved from the New York State ECLRS, addresses were matched with tax parcel data to determine whether the household was connected to public sewer, and then geocoded to sewershed geographies before aggregation into daily numbers. Test positivity was defined as the number of COVID-19 cases divided by the number of COVID-19 tests conducted. The number of active cases was estimated as each COVID-19 case lasting 10 days from diagnosis. We utilized seven-day averages for test positivity and incidence, but not for active cases. We estimated simple Pearson correlations between levels of SARS-CoV-2 RNA in wastewater and measures of incidence, active cases, and test positivity.

#### Sewershed population estimates

We calculated estimates for the 2020 population within each sewershed using R statistical software version 4.0.0.^36^ We first estimated the 2010 population for each sewershed using an overlay of 2010 US census blocks on top of the sewershed boundaries. We calculated the proportion of the area for partial block overlap and then assigned a proportional 2010 decennial population of the block to the sewershed assuming equal distribution of the population in the blocks. We then aggregated the apportioned values to get a total population estimate for the sewershed. We repeated this procedure using 2010 decennial population data for the block group and 2018 American Community Survey (ACS) data for the block group to get 2010 and 2018 population by sewershed based on block groups. We used these values to estimate the rate of population change per sewershed using equation 1. We then applied this average annual change to the sewershed population based on the block data from 2010 and estimated the population after ten years of growth using equation 2 to calculate 2020 population estimates. The “tidycensus” R package provided population estimates^37^ and the “tigris” package provided geometry data.^38^

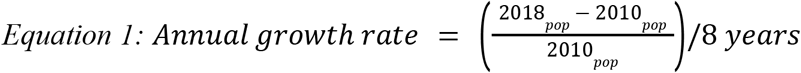

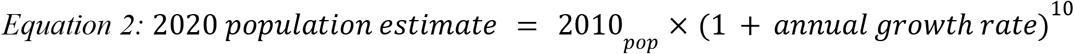

#### Freedom from transmission estimates

In order to estimate the limits of detection of wastewater surveillance we first categorized wastewater results into not detected, detected but below the limits of quantification, and quantifiable. We then examined the reported cases and test positivity in the sewershed as a function of each of these categories. We calculated sensitivity to confirm absence of transmission as the proportion of sewersheds reporting < 1 daily incident COVID-19 case per 10,000 population at the time of the wastewater sample. Following the calculated limits of detection we estimated the sensitivity of the wastewater surveillance platform to detect SARS-CoV-2 transmission as a function of the calculated limits of detection, a correction factor for the population size of the sewershed, and the proportion of houses within the sewershed who are connected to the sewer system using equations 3 and 4. The population correction factor in these equations refers to how much larger the sewershed population is than 10,000 population, being one if smaller than 10,000 and otherwise the value of the sewershed population divided by 10,000.

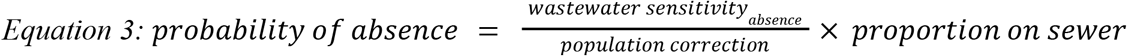

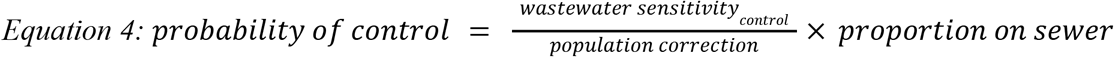

## Supplemental figures

**Supplemental Figure 1:**
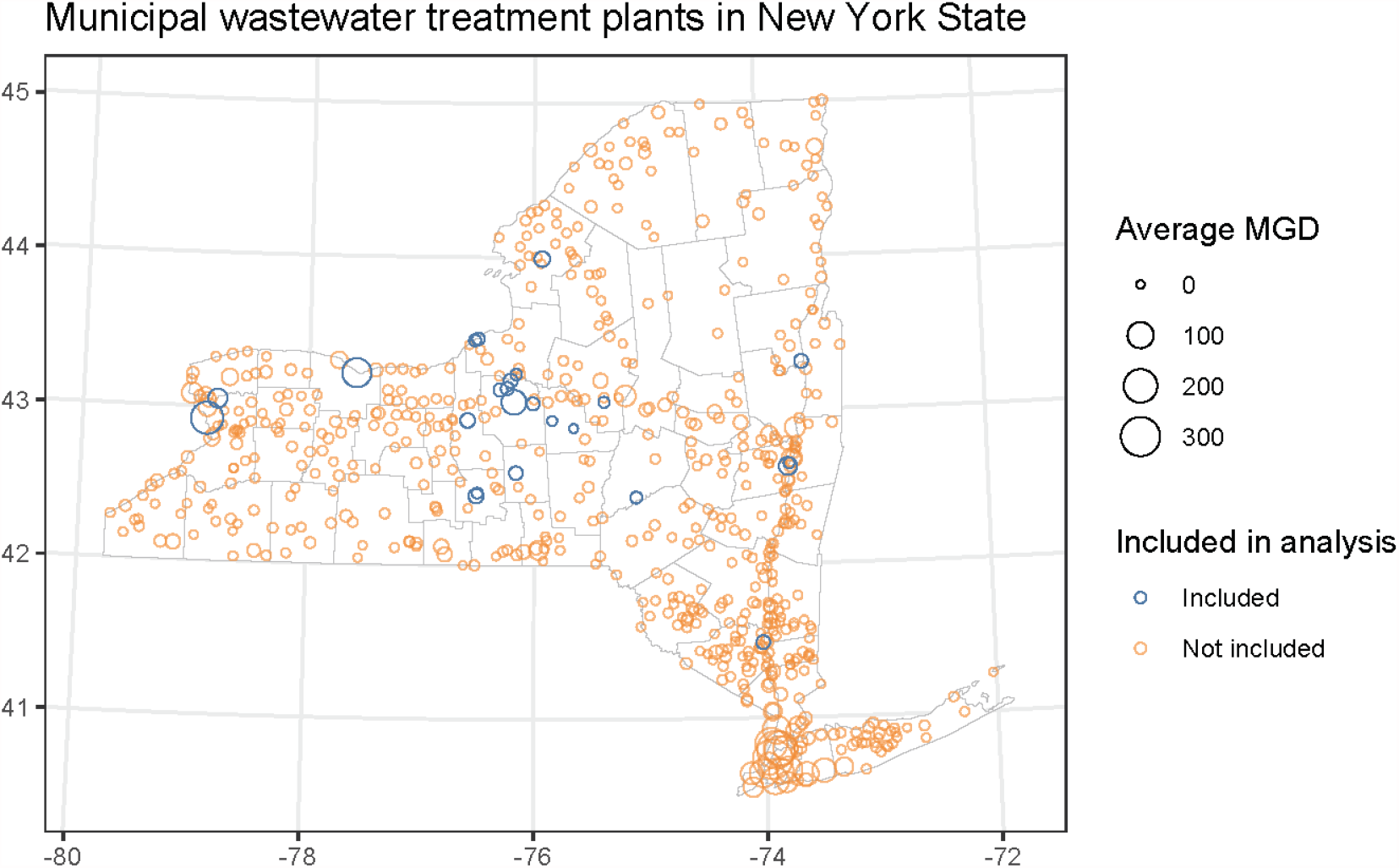
Municipal wastewater treatment plants in New York State. MGD = Million gallons per day.

**Supplemental figure 2:**
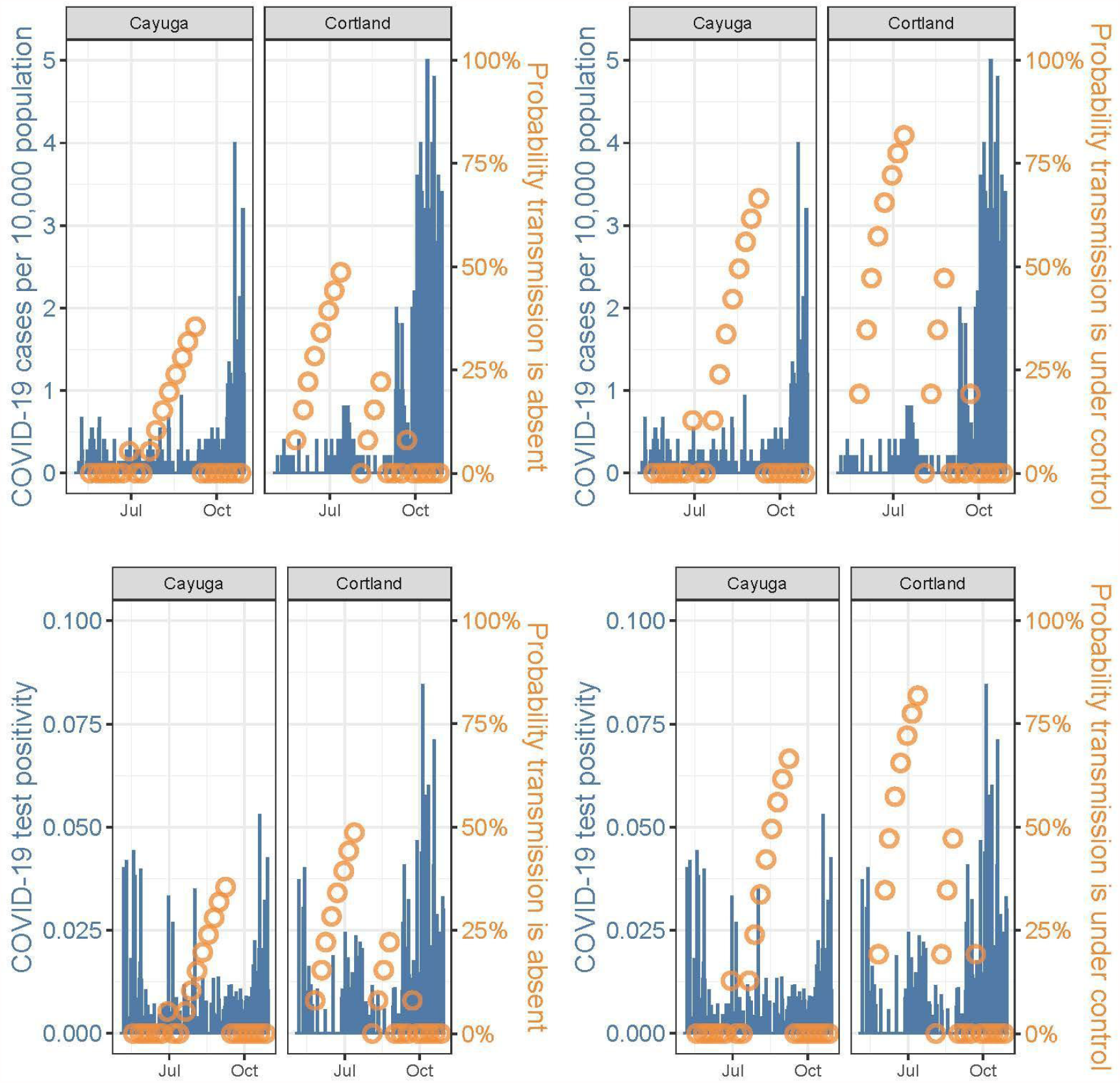
Probability that coronavirus transmission was absent or under control with repeated non-detection of SARS-CoV-2 RNA in wastewater over time alongside incidence of COVID-19 cases and test positivity in the county.

**Supplemental Figure 3:**
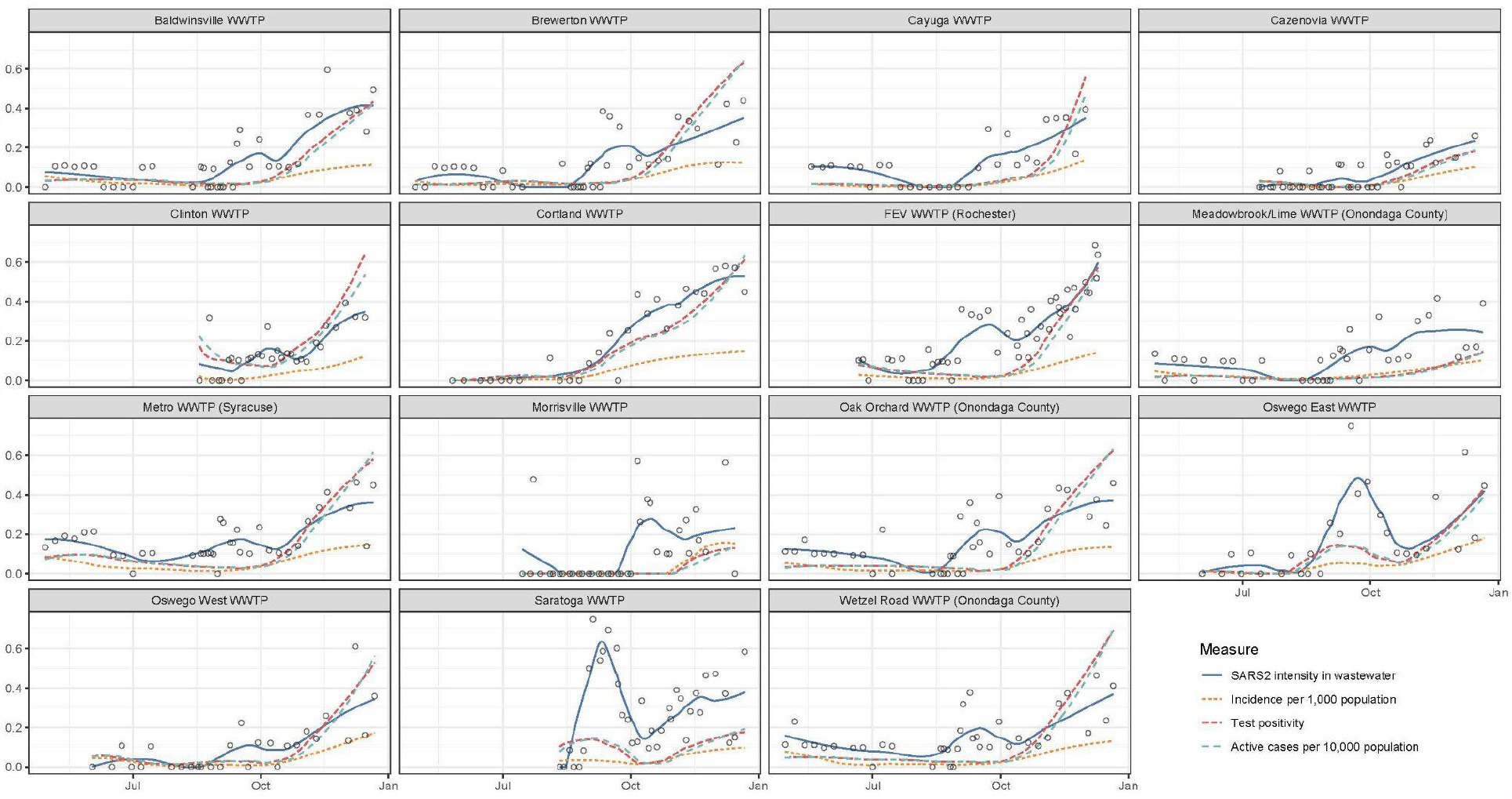
Trends in SARS-CoV-2 transmission in various communities throughout upstate New York in 2020 as measured by the intensity of SARS-CoV-2 RNA in wastewater, COVID-19 incidence, COVID-19 test positivity, and active COVID-19 cases. Points represent measured SARS-CoV-2 intensity, while lines are smoothed with estimates.

**Supplemental figure 4:**
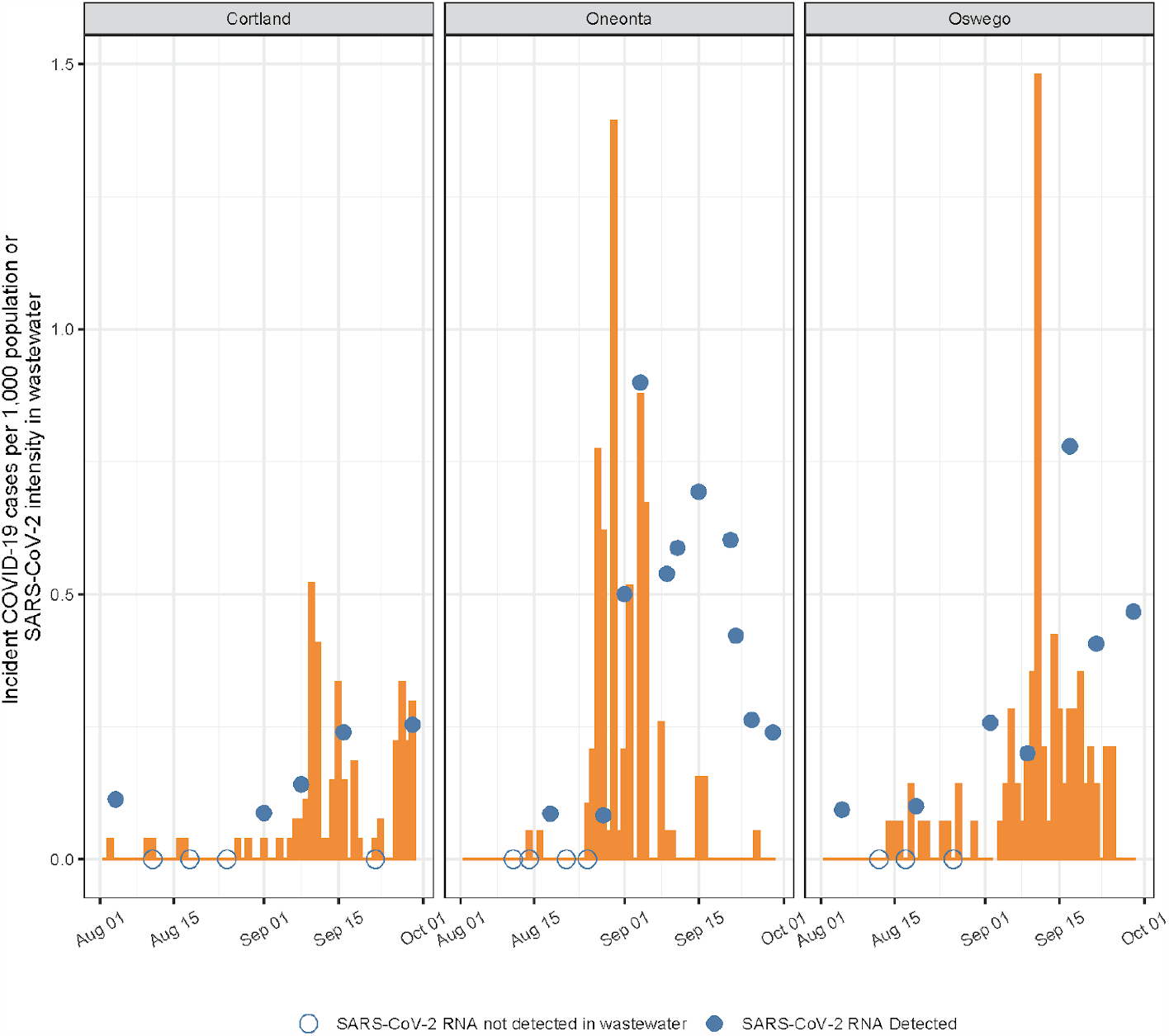
COVID-19 cases (bars) and wastewater results (circles) from three communities with COVID-19 outbreaks in the fall of 2020. Early indication of increasing transmission was provided by non-quantifiable but detected levels of SARS-CoV-2 RNA in wastewater.

## References and Notes

1. Thacker, S. B. & Berkelman, R. L. Public health surveillance in the United States. Epidemiologic Reviews 10, 164–190 (1988).

2. Rushton, S. Global Health Security: Security for whom? Security from what? Political Studies 59, 779–796 (2011).

3. Aldis, W. Health security as a public health concept: a critical analysis. Health Policy and Planning 23, 369–375 (2008).

4. Abat, C., Chaudet, H., Rolain, J.-M., Colson, P. & Raoult, D. Traditional and syndromic surveillance of infectious diseases and pathogens. International Journal of Infectious Diseases 48, 22–28 (2016).

5. Dato, V., Wagner, M. M. & Fapohunda, A. How Outbreaks of Infectious Disease are Detected: A Review of Surveillance Systems and Outbreaks. Public Health Rep 119, 464–471 (2004).

6. Chan, J. F.-W. et al. A familial cluster of pneumonia associated with the 2019 novel coronavirus indicating person-to-person transmission: a study of a family cluster. The Lancet 395, 514–523 (2020).

7. Shear, M. D. et al. The Lost Month: How a Failure to Test Blinded the U.S. to Covid-19. The New York Times (2020).

8. Dowell, S. F., Blazes, D. & Desmond-Hellmann, S. Four steps to precision public health. Nature 540, 189–191 (2016).

9. Khoury, M. J., Iademarco, M. F. & Riley, W. T. Precision Public Health for the Era of Precision Medicine. American Journal of Preventive Medicine 50, 398–401 (2016).

10. Cuomo, A. No. 202.4: Continuing Temporary Suspension and Modification of Laws Relating to the Disaster Emergency. https://www.governor.ny.gov/news/no-2024-continuing-temporary-suspension-and-modificat ion-laws-relating-disaster-emergency (2020).

11. Larsen, D. A. & Wigginton, K. R. Tracking COVID-19 with wastewater. Nat Biotechnol (2020) doi:10.1038/s41587-020-0690-1.

12. Chen, Y. et al. The presence of SARS-CoV-2 RNA in the feces of COVID-19 patients. Journal of Medical Virology 92, 833–840 (2020).

13. Wang, W. et al. Detection of SARS-CoV-2 in Different Types of Clinical Specimens. JAMA - Journal of the American Medical Association 323, 1843–1844 (2020).

14. Xu, Y. et al. Characteristics of pediatric SARS-CoV-2 infection and potential evidence for persistent fecal viral shedding. Nature Medicine 26, 502–505 (2020).

15. Medema, G., Heijnen, L., Elsinga, G., Italiaander, R. & Brouwer, A. Presence of SARS-Coronavirus-2 RNA in Sewage and Correlation with Reported COVID-19 Prevalence in the Early Stage of the Epidemic in The Netherlands. Environ. Sci. Technol. Lett. 7, 511–516 (2020).

16. Peccia, J. et al. Measurement of SARS-CoV-2 RNA in wastewater tracks community infection dynamics. Nat Biotechnol (2020) doi:10.1038/s41587-020-0684-z.

17. Wurtzer, S. et al. Evaluation of lockdown effect on SARS-CoV-2 dynamics through viral genome quantification in waste water, Greater Paris, France, 5 March to 23 April 2020. Eurosurveillance 25, (2020).

18. Naughton, C. C. et al. Show us the Data: Global COVID-19 Wastewater Monitoring Efforts, Equity, and Gaps. http://medrxiv.org/lookup/doi/10.1101/2021.03.14.21253564 (2021) xdoi:10.1101/2021.03.14.21253564.

19. Gray, J. D. A. The isolation of B. Paratyphosus B from sewage. Br Med J 1, 142–144 (1929).

20. Kelly, S. M., Clark, M. E. & Coleman, M. B. Demonstration of Infectious Agents in Sewage. Am J Public Health Nations Health 45, 1438–1446 (1955).

21. Moore, B. The detection of enteric carriers in towns by means of sewage examination. J R Sanit Inst 71, 57–60 (1951).

22. Vaccines and Biologicals & World Health Organization. Guidelines for environmental surveillance of poliovirus circulation. (2003).

23. Asghar, H. et al. Environmental surveillance for polioviruses in the global polio eradication initiative. Journal of Infectious Diseases 210, S294–S303 (2014).

24. Brouwer, A. F. et al. Epidemiology of the silent polio outbreak in Rahat, Israel, based on modeling of environmental surveillance data. Proceedings of the National Academy of Sciences of the United States of America 115, E10625–E10633 (2018).

25. Cameron, A. R. & Baldock, F. C. A new probability formula for surveys to substantiate freedom from disease. Preventive Veterinary Medicine 34, 1–17 (1998).

26. Green, H. et al. Quantification of SARS-CoV-2 and cross-assembly phage (crAssphage) from wastewater to monitor coronavirus transmission within communities. medRxiv 2, 2020.05.21.20109181-2020.05.21.20109181 (2020).

27. Black, J. et al. Epidemiological evaluation of sewage surveillance as a tool to detect the presence of COVID-19 cases in a low case load setting. Science of The Total Environment 147469 (2021) doi:10.1016/j.scitotenv.2021.147469.

28. Russell, T. W., Hellewell, J., Abbott, S., van Zandvoort, K. & Ratnayake, R. Using a delay-adjusted case fatality ratio to estimate under-reporting. 6 (2020).

29. Lieberman-Cribbin, W., Tuminello, S., Flores, R. M. & Taioli, E. Disparities in COVID-19 Testing and Positivity in New York City. American Journal of Preventive Medicine 59, 326–332.

30. Sun, K. et al. Transmission heterogeneities, kinetics, and controllability of SARS-CoV-2. Science 371, (2021).

31. Woolhouse, M. E. et al. Heterogeneities in the transmission of infectious agents: implications for the design of control programs. Proceedings of the National Academy of Sciences of the United States of America 94, 338–342 (1997).

32. Hellewell, J. et al. Feasibility of controlling COVID-19 outbreaks by isolation of cases and contacts. The Lancet Global Health 8, e488–e496 (2020).

33. Aleta, A. et al. Modelling the impact of testing, contact tracing and household quarantine on second waves of COVID-19. Nature Human Behaviour 4, 964–971 (2020).

34. Thompson, C. N. et al. COVID-19 Outbreak — New York City, February 29–June 1, 2020. MMWR Morb. Mortal. Wkly. Rep. 69, 1725–1729 (2020).

35. Electronic Clinical Laboratory Reporting System - ECLRS. https://www.health.ny.gov/professionals/reportable_diseases/eclrs/.

36. R Core Development Team. R: A Language and Environment for Statistical Computing. http://www.R-project.org/ (2010).

37. Walker, K. & Herman, M. tidycensus: Load US Census Boundary and Attribute Data as ‘tidyverse’ and ‘sf ‘-Ready Data Frames. (2021).

38. Walker, K. tigris: Load Census TIGER/Line Shapefiles. (2020).

39. Kilaru, P., Larsen, D. & Monk, D. Design and utilization of homemade wastewater samplers during the COVID-19 pandemic. https://engrxiv.org/frbuk/ (2020) xdoi:10.31224/osf.io/frbuk.

40. Sikorski, M. J. & Levine, M. M. Reviving the “Moore Swab”: a Classic Environmental Surveillance Tool Involving Filtration of Flowing Surface Water and Sewage Water To Recover Typhoidal Salmonella Bacteria. Appl. Environ. Microbiol. 86, (2020).

41. Feng, S. et al. Evaluation of sampling frequency and normalization of SARS-CoV-2 wastewater concentrations for capturing COVID-19 burdens in the community. http://medrxiv.org/lookup/doi/10.1101/2021.02.17.21251867 (2021) xdoi:10.1101/2021.02.17.21251867.

42. Williams Walsh, M. Virus Did Not Bring Financial Rout That Many States Feared. The New York Times (2021).

